# Differential expression analyses on aortic tissue reveal novel genes and pathways associated with abdominal aortic aneurysm onset and progression

**DOI:** 10.1101/2024.02.26.24303384

**Authors:** Gerard Temprano-Sagrera, Begoña Soto, Jaume Dilmé, Olga Peypoch, Laura Calsina Juscafresa, David Davtian, Lluís Nieto, Andrew Brown, José Román Escudero, Ana Viñuela, Mercedes Camacho, Maria Sabater-Lleal

## Abstract

**Background:** Abdominal aortic aneurysms (AAA) are focal dilatations of the abdominal aorta. They are normally asymptomatic and progressively expand, increasing their risk of rupture. Rupture of an AAA is associated with high mortality rates, but the mechanisms underlying the initiation, expansion and rupture of AAA are not yet fully understood. This study aims to characterize and identify new genes associated with the pathophysiology of AAA through differential expression analyses between dilated and non-dilated aortic tissue samples, and between AAA of different diameters. Our study used RNA-seq data on 140 samples, becoming the largest RNA-seq dataset for differential expression studies of AAA.

**Results:** We identified 7,454 differentially expressed genes (DEGs) between AAA and controls, 2,851 of which were new compared to previous microarray studies. Notably, a novel cluster on adenosine triphosphate synthesis regulation emerged as strongly associated with AAA. Additionally, exploring AAA of different diameters identified eight genes (*EXTL3*, *ZFR*, *DUSP8*, *DISP1*, *USP33*, *VPS37C*, *ZNF784*, *RFX1*) that overlapped with the DEGs between AAA and controls, implying roles in both disease onset and progression. Seven genes (*SPP1*, *FHL1*, *GNAS*, *MORF4L2*, *HMGN1*, *ARL1*, *RNASE4*) with differential splicing patterns were also DEGs between AAA and controls, suggesting that splicing differences contribute to the observed expression changes and the disease development.

**Conclusions:** This study identified new genes and pathways associated with AAA onset and progression and validated previous relevant roles of inflammation and intracellular calcium regulation. These findings provide insights into the complex mechanisms underlying AAA and indicate potential targets to limit AAA progression and mortality risk.

## BACKGROUND

Abdominal aortic aneurysms (AAA) are characterized by a local dilation of the infrarenal abdominal aorta to about 1.5 times the normal adjacent aortic diameter or more than 3 cm in maximum diameter[1]. AAA is accompanied by chronic inflammation, apoptosis of vascular smooth muscle cells and neovascularization[2,3]. Additionally, extracellular matrix degradation, microcalcification, and oxidative stress contribute to the degeneration of the aorta[1,2]. The disease is progressive, and most aneurysms develop without causing symptoms[1]. However, in the event of AAA rupture, mortality rates can reach 80 %[4]. The only effective treatment currently available for AAA is aortic tissue repair, either through open surgery or endovascular repair[1,5].

Some risk factors are known for developing AAA, including age, male sex, smoking, and family history of AAA[1]. Smoking, in addition, is also known to increase the risk of rupture[6]. Additionally, recent genomic studies have revealed 121 loci associated with risk of developing AAA, contributing to the knowledge of the possible pathways leading to this disease[7]. However, there is still an insufficient understanding of the clear mechanisms that underlie the initiation, propagation, and rupture of AAA.

The study of gene expression, known as transcriptomics, is a valuable tool for understanding human disease and revealing new therapeutic targets [8,9]. Several studies have been performed to study the differentially expressed genes (DEGs) between dilated aortic tissue and non-dilated control aorta using microarray technology, detecting DEGs especially associated with the immune and inflammatory responses, extracellular matrix remodeling and angiogenesis[10–15]. In the present study, we performed RNA sequencing (RNAseq) of the complete transcriptome in 140 abdominal aortic tissue samples (96 dilated aortas and 44 control aortas from deceased donors) from the Triple A Barcelona Study (TABS) cohort, to identify new DEGs and pathways associated with the pathophysiology of AAA initiation and progression, allowing for a more comprehensive analysis of the transcriptome and becoming the largest RNAseq dataset for AAA tissue. Additionally, we aimed to investigate the differences in alternative splicing patterns in the context of AAA, and the role of genetic variants in gene expression in AAA tissue. The study design is described in **Figure 1**.

**Figure 1.**
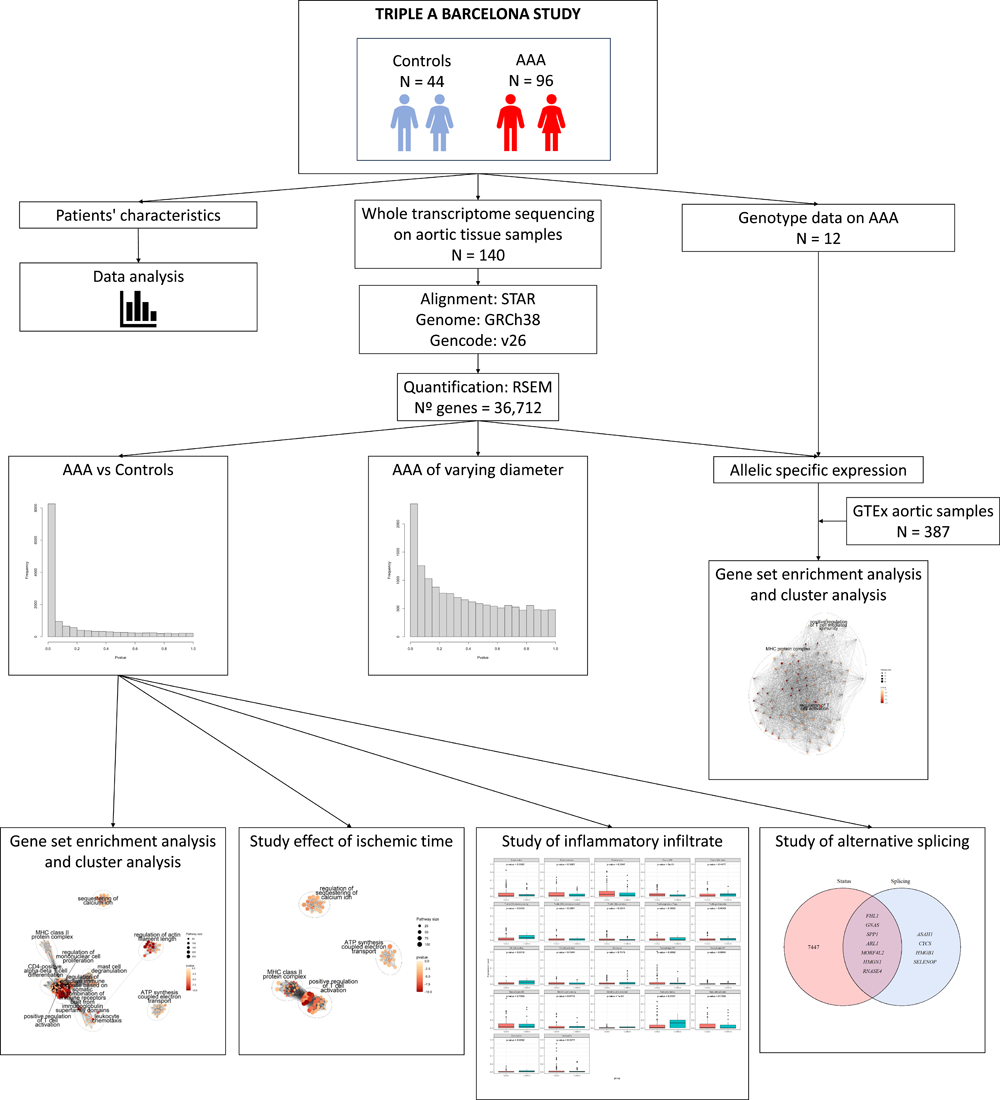
Study design flowchart.

## RESULTS

### Participants characteristics

**Table 1** shows the participants demographic and clinical data. Aortic tissue samples from 96 AAA patients and 44 controls from the TABS cohort were used for RNAseq analysis.

**Table 1:** Characteristics of study participants.

|  | **Controls**<br>**(N = 44)** | **AAA**<br>**(N = 96)** | **P-value** | **Missing**<br>**values (%)** |
| --- | --- | --- | --- | --- |
| **Age (Years)** | 61.66 (21-82) | 70.38 (53-87) | 0.0006 | 0 (0) |
| **Sex (Male)** | 21 (47.73) | 92 (95.83) | 1.00E-10 | 0 (0) |
| **Smoking (Current)** | 8 (21.62) | 27 (32.14) | 0.3378 | 19 (13.57) |
| **Smoking (Past)** | 3 (8.11) | 42 (50) | 2.81E-05 | 19 (13.57) |
| **Aortic Diameter (mm)** | NA | 65.57 (38-100.12) | - | 0 (0) |
| **Hypertension (Yes)** | 15 (40.54) | 54 (64.29) | 0.0256 | 19 (13.57) |
| **Dyslipidemia (Yes)** | 10 (27.03) | 46 (54.76) | 0.0087 | 19 (13.57) |
| **Diabetes mellitus (Yes)** | 5 (13.51) | 12 (14.29) | 1 | 19 (13.57) |
| **Peripheral arterial disease (Yes)** | NA | 23 (27.71) | - | 57 (59.38) |
| **Other aneurysms (Yes)** | NA | 25 (29.76) | - | 12 (12.5) |
| **Cerebrovascular Disease (Yes)** | 3 (8.11) | 36 (43.9) | 0.0003 | 21 (15) |
| **Cardiovascular Disease (Yes)** | 1 (3.34) | 16 (19.05) | 0.0846 | 27 (19.29) |
| **Chronic obstructive pulmonary disease (Yes)** | 1 (2.7) | 15 (17.86) | 0.0481 | 19 (13.57) |
| Chronic kidney disease (Yes) | 2 (5.4) | 17 (20.24) | 0.0726 | 19 (13.57) |

Continuous variables are presented as mean (range), and categorical variables are presented as %. Two-sample t-tests and chi-squared tests were used to compare the means of continuous phenotypes and the distribution of categorical phenotypes, between AAA and control groups, respectively. Missing values were excluded from the calculations of each variable. Hypertension was defined based on clinical history and the use of antihypertensive medication. Dyslipidemia was diagnosed through clinical history and the use of hypolipidemic medication. Diabetes mellitus was identified by clinical history and the use of insulin or oral hypoglycemic medications, without differentiation between type 1 or type 2. Peripheral arterial disease was assessed based on clinical symptoms and clinical history. Other aneurysms included thoracic and visceral aortic aneurysms, iliac artery aneurysms, and popliteal artery aneurysms, and were diagnosed using computed tomography or ultrasound. Cerebrovascular diseases were determined by a history of transient ischemic attack or stroke. Cardiovascular diseases were assessed by history of acute myocardial infarction or angina pectoris, or admission with clinical symptoms, electrocardiogram changes, or a positive enzymatic curve diagnosed by a cardiologist. Chronic obstructive pulmonary disease was identified based on clinical history. Chronic kidney disease was assessed by clinical history.

We examined demographic and clinical variables between AAA and controls to determine whether these might influence our expression levels differences. We found significant differences in sex (Additional file 1: Figure S1A), age (Additional file 1: Figure S1B), smoking status, and the prevalence of hypertension, dyslipidemia, cerebrovascular disease, and chronic obstructive pulmonary disease between AAA patients and controls. In relation to smoking status (N = 122), which is a known risk factor for AAA development and rupture, 70.3 % of controls were never smokers, compared to 17.86 % of AAA patients. Among current and past smokers, there were also differences, with 50 % of former smokers and 32.14 % of current smokers in AAA, compared to only 8.11 % and 21.62 % respectively, in the controls (Additional file 1: Figure S3C). We adjusted our regression analyses for age and sex, acknowledging their potential influence on expression level differences.

### Differential expression analyses between AAA and controls

The analysis of differential expression between aortic samples from 96 AAA patients and 44 controls revealed 7,454 genes displaying significant differences in expression (adjusted p-value < 0.05) (Additional file 1: Figure S2A and Additional file 2: Table S1). This number exceeds by 26.57 % the previous DEGs identified in comparable analyses using microarray technologies[10–15]. Using GO and KEGG enrichment analyses, we found a total of 1,152 and 89 enriched terms, respectively (Additional file 1: Figure S3A and Figure S4A). The complete results of the enriched terms for GO and KEGG are shown in Additional files 3 and 4: Tables S2 and S3. To better characterize the biological processes associated with the DEGs in the enrichment analysis, we performed a cluster analysis of GO pathways. We found that most of the DEGs were associated with the immune system: regulation of mononuclear cell proliferation, leukocyte chemotaxis, regulation of adaptive immune response based on somatic recombination of immune receptors built from immunoglobulin superfamily domains, mast cell degranulation, major histocompatibility complex (MHC) class II protein complex, positive regulation of T cell activation, and CD4-positive alpha-beta T cell differentiation (Figure 2A). Other represented metabolic pathways were related to sequestering of calcium ion, regulation of actin filament length and adenosine triphosphate (ATP) synthesis coupled electron transport (Figure 2A). While these analyses corroborated previous associations with inflammatory, actin filament regulation and intracellular calcium regulation processes, the ATP synthesis regulation pathway was a new differential expressed pathway in AAA tissue.

**Figure 2.**
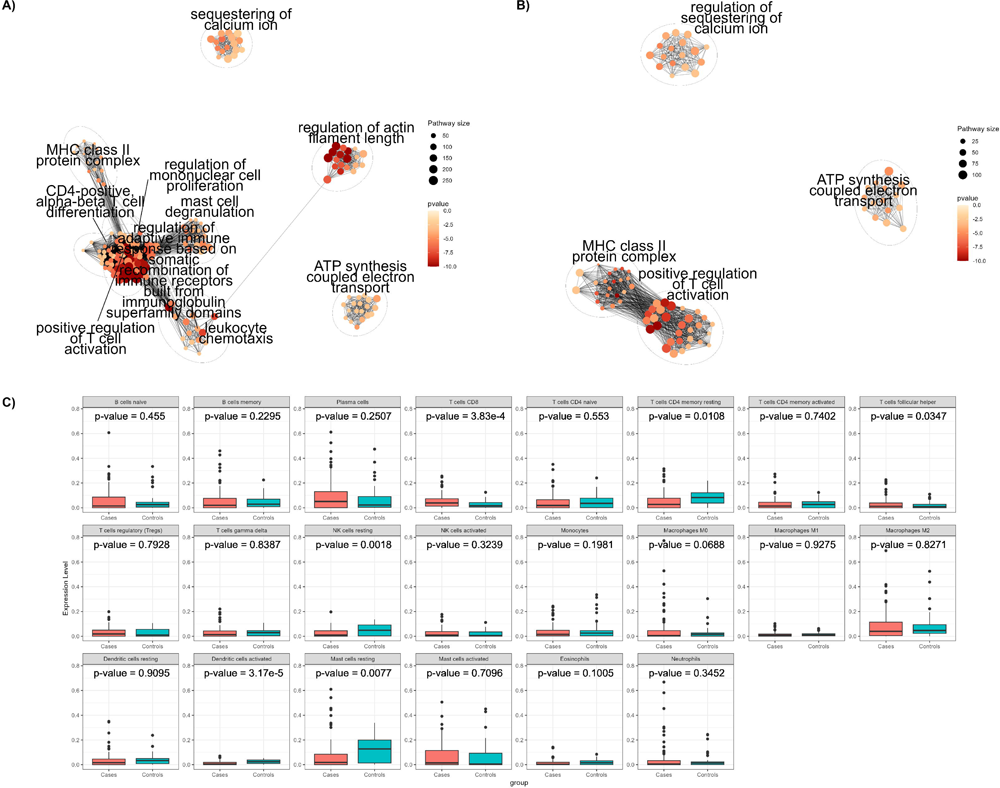
Hierarchical clustering analysis results with all the DEGs between AAA and controls (A) and after removing DEG by ischemic time (B). C) Comparison of the proportion of inflammatory cells between AAA and controls using CIBERSORTx.

Control aortic samples from deceased organ donors have been used by us and others [10–13,15]. To account for differences in gene expression between AAA and control tissue that could be attributed to ischemic time (time between the donor’s death and sample collection when blood flow is interrupted), we removed 10,737 DEGs associated to ischemic time in the GTEx aorta samples (N = 387)[16] from the total DEGs found between AAA and controls, leaving 3,002 DEGs (Additional file 1: Figure S2B and Additional file 5: Table S4).

We then performed a new enrichment analysis and identified 424 enriched GO terms and 65 KEGG pathways (Additional file 1: Figure S3B and Figure S4B) (Complete results are available at Additional files 6 and 7: Tables S5 and S6, respectively), which represented removal of 728 and 24 pathways susceptible of being caused by ischemic time, respectively. Cluster analysis of GO enriched terms confirmed identified clusters related to the regulation of calcium ion retention, ATP synthesis coupled electron transport, and immune response centered on T cell activation (MHC class II protein complex, positive regulation of T cell activation). On the other hand, other clusters also associated with the immune system were no longer represented (regulation of mononuclear cell proliferation, leukocyte chemotaxis regulation of adaptive immune response based on somatic recombination of immune receptors built from immunoglobulin superfamily domains, CD4-positive, alpha-beta T cell differentiation, mast cell degranulation) together with the regulation of actin filament length (Figure 2B). By accounting for genes whose expression was altered by ischemic time, we identified a set of genes that are less likely to be affected by the experimental limitations of these types of studies.

Vascular inflammation has been previously associated to AAA development and progression[17,18]. Even in our most stringent analysis, which removed pathways possibly caused by ischemic time, there was a notable enrichment of pathways associated with the immune response. Consequently, we decided to investigate the influence of the inflammatory infiltrate on AAA by comparing the abundance of 22 immune cell types from gene expression profiles between our AAA and control samples. After correcting for multiple testing, we found significant differences on the abundances of CD8 T-cells, natural killer (NK) resting cells, and dendritic activated cells (Figure 2C). AAA samples had a higher proportion of CD8 T-cells, while controls had more NK resting cells and dendritic activated cells. Table 2 provides a summary of the three cell populations with the highest and lowest presence in each group. The findings support the previously reported involvement of CD8 T-cells and NK cells in AAA.[17,18] The lower levels of dendritic cells detected in AAA were unexpected, as previous studies found significantly higher levels of dendritic cells in AAA samples compared to controls[19].

**Table 2:** Summary of the three most and least prevalent cell populations in AAA and controls.

| Cell Populations (AAA) | Percentage (%) | Cell Populations (Controls) | Percentage (%) |
| --- | --- | --- | --- |
| Plasma B cells | 9.46 | Resting mast cells | 12.34 |
| M2 macrophages | 8.68 | Resting memory CD4 T cells | 8.31 |
| Resting mast cells | 6.99 | M2 Macrophages | 8.23 |
| M1 macrophages | 1.43 | Eosinophils | 2.03 |
| Eosinophils | 1.37 | T cell follicular helper cells | 1.94 |
| Activated dendritic cells | 1.13 | M1 Macrophages | 1.45 |

### Study of alternative splicing

The relevance of alternative splicing in the development of diseases, such as cancer, neurological, and cardiovascular diseases has been well-established for years.[20] However, capturing the complexity of alternative splicing has been challenging. With the recent improvements in sequencing techniques it is now possible to study alternative splicing in more depth.[20] We investigated alternative splicing patterns between the AAA and control groups to identify specific splicing patterns associated with AAA. We identified 15 significant alternative splicing events on eleven unique genes (FHL1, GNAS, ASAH1, SPP1, ARL1, MORF4L2, CYCS, HMGB1, HMGN1, SELENOP, RNASE4) between AAA and controls. The analysis revealed that, as anticipated, the number of altered alternative first exon events was more represented than any other splice event among AAA and control samples (Figure 3A), consistent with previous work suggesting that it is the most frequent splice event in the human genome[21] (Complete results are available in Additional file 8: Table S7). A functional enrichment analysis was conducted on the eleven genes with significant alternative splicing events, but no significantly enriched metabolic pathways were found. Interestingly, seven out of the eleven genes were also differentially expressed between AAA and controls, suggesting that the expression of specific splicing variants could be altered in AAA (Figure 3B).

**Figure 3.**
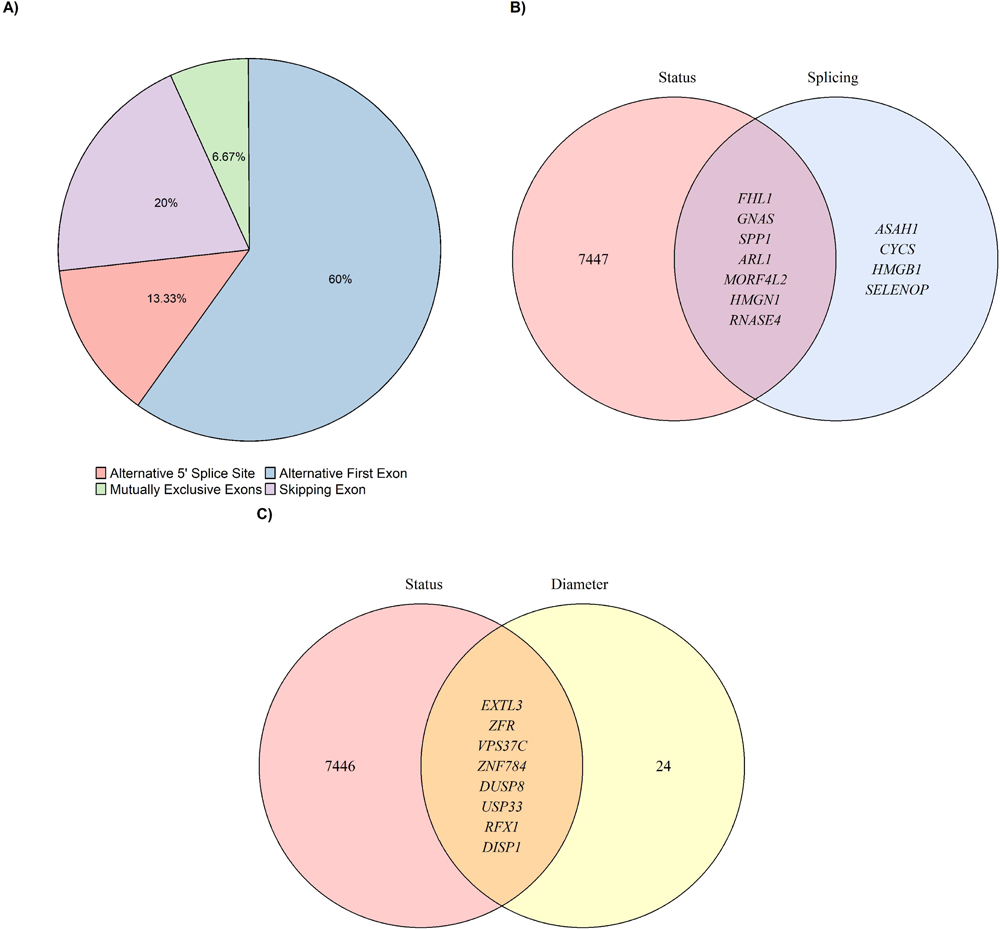
A) Significant alternative splicing types identified between AAA and controls. B) Venn diagram showing the overlap between DEGs in AAA and controls and genes with differential alternative splicing patterns in AAA and controls. C) Venn diagram showing the overlap between DEGs in AAA and controls and DEGs by AAA diameter.

### Differential expression analysis by aortic diameter

The diameter of AAA is a significant risk factor for rupture. We analyzed the DEGs by diameter to identify alterations in gene expression throughout the progression of the disease. We observed a total of 32 DEGs among aneurysms of varying diameters (N = 84), although no enriched pathways were identified (Additional file 1: Figure S2C and Additional file 9: Table S8). Of the 32 DEGs by diameter, eight were also DEGs between AAA and controls (Figure 3C), suggesting that these eight genes are relevant for disease formation and also during disease progression.

### Allelic specific expression

We investigated the potential effect of AAA-associated genetic variants on gene expression in diseased tissue by studying allele specific expression in twelve AAA samples with available genetic data. On average, we identified 529 genes with significant allele specific expression (adjusted p-value < 0.05) in the twelve AAA samples. Among these genes, 90 exhibited significant allele specific expression in more than five of the twelve AAA samples. Additionally, to determine whether these associations were related to AAA or were characteristic of the aortic tissue, we compared allele specific expression patterns between our AAA samples and 387 GTEx aortic samples, used as controls. The comparison between AAA and control samples identified 1,815 genes with significant differences in the allele specific expression patterns (adjusted p-value < 0.05) (Figure 4A). An enrichment analysis on GO terms for these 1,815 genes revealed 91 enriched pathways. The posterior cluster analysis revealed three clusters strongly related to the immune system: MHC protein complex, positive regulation of T cell-mediated cytotoxicity, and regulation of T cell activation (Figure 4B). The association between the immune system and AAA was also observed in the differential expression analysis between AAA samples and controls, validating the robustness of these results.

**Figure 4.**
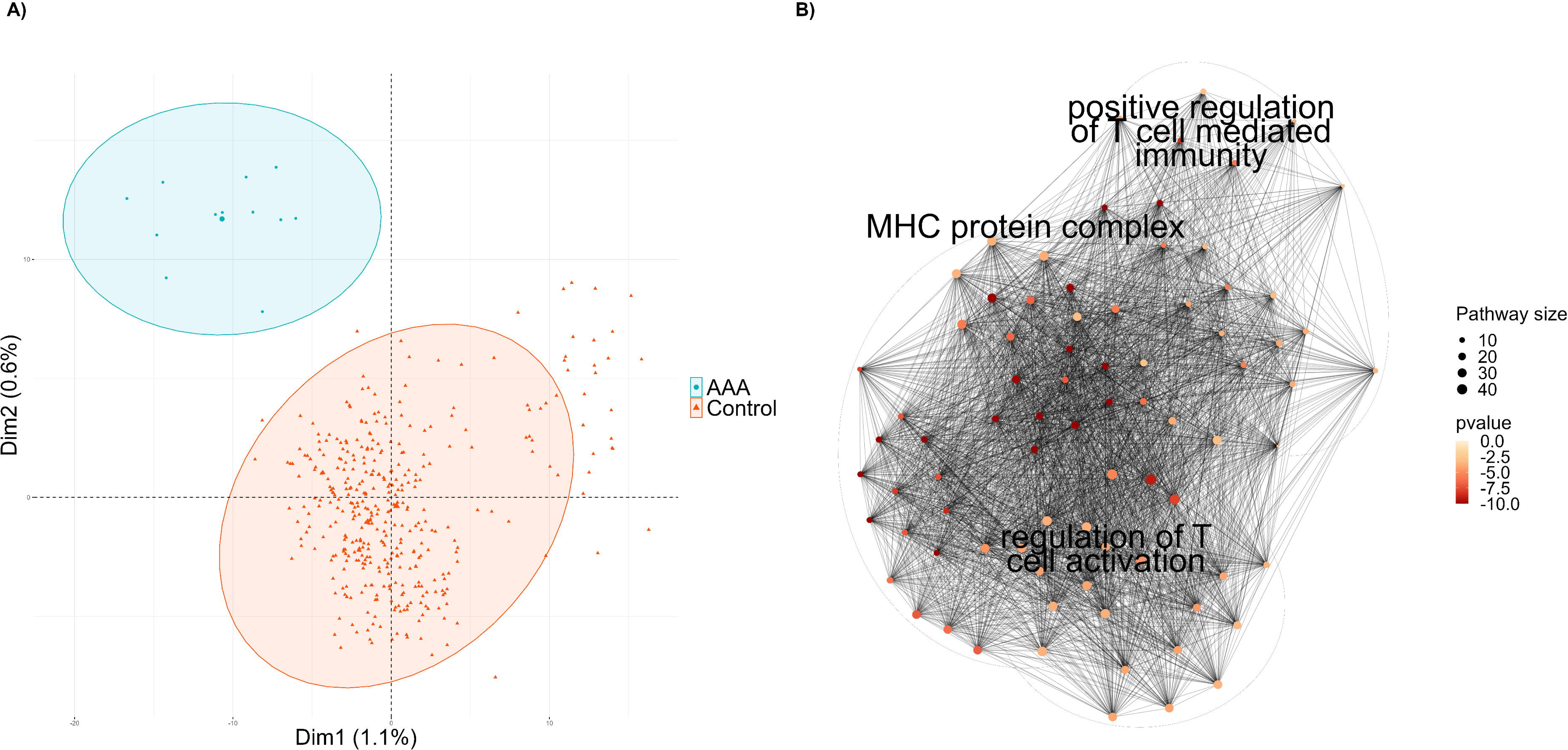
A) Results of PC analysis clustering between AAA and controls. B) Hierarchical clustering analysis results with genes with different allelic specific patterns between AAA and controls.

Finally, among the 90 genes that exhibited significant allele specific expression in more than five AAA samples, we selected those that also showed significant differential allelic specific expression analysis between AAA and GTEx control samples, and those present in loci identified in the largest genome-wide association study (GWAS) on AAA risk[7], in order to identify haplotypes associated with AAA risk. Among the selected genes, SNURF was the only gene that also presented differential allele specific expression patterns between AAA and control tissues, and SPP1 and THBS2 were prioritized based on their presence in a locus identified in the previous GWAS on AAA. This allowed to hypothesize that the presence of particular genetic haplotypes in these three genes determined their differential expression associated with risk of AAA.

## DISCUSSION

This study analyzes differential expression between AAA aortic tissue samples and control aortic samples using whole transcriptome data obtained through RNAseq. In addition, we studied the effect of ischemic time on gene expression, to obtain a more credible list of genes associated with AAA development. Using our RNAseq data, which provides superior alternative splicing analysis compared to microarrays[22], we conducted a novel exploration of alternative splicing between AAA and control samples, to identify potential causes of the observed differential expression. Furthermore, we analyzed the differential expression between AAA of different diameters to study the genes altered during disease progression. Finally, we analyzed allele specific expression to gain insights into how genetic variants impact expression in the diseased tissue.

### Study of ischemic time-independent pathways involved in AAA development

Clustering analysis with the enriched pathways after accounting for the ischemic time effect revealed a strong association with MHC class II protein complex, positive regulation of T-cells, and intracellular calcium ion regulation. Additionally, we have for the first time identified the regulation of the ATP synthesis pathway in a differential expression analysis of aortic samples. While the detection of the ATP synthesis regulatory pathway is novel, it is in line with previous work associating mitochondrial disfunction and AAA [23,24]. On the other hand, previous differential expression studies in microarrays between AAA and control aortic tissue have consistently found associations of immune system pathways with AAA.[10–13,15] Our results confirm these associations and demonstrate that these are independent of the ischemic time, which is a confounder factor in most studies using donor samples. Finally, the regulation of intracellular calcium was previously detected in one study of differential expression between dilated and non-dilated aortic samples[11]. Our analyses confirm that this association is independent of ischemic time.

We identified for the first time several enriched signaling pathways, with a large presence of genes that code for subunits of complexes I (NADH ubiquinone oxidoreductase)), III (Ubiquinol-cytochrome c reductase) and IV (cytochrome c oxidase)) of the electron transport chain. Our results indicate that 88 % (22 / 25) of the DEGs coding for the subunits of the complexes that form the electron transport chain are expressed to a lower extent in AAA, suggesting a lower synthesis of ATP in AAA tissue. Mitochondrial dysfunction has previously been studied in the development of AAA[23,24] and other cardiovascular diseases[25] due to its key role in some of the cellular alterations characteristic of cardiovascular diseases, including excessive production of reactive oxygen species, energy depletion, endoplasmic reticulum stress, and activation of apoptosis. However, this is the first time that these metabolic pathways have been characterized in a DEGs study of AAA, confirming the role of mitochondrial dysfunction on AAA.

Among the genes present in all the enriched signaling pathways related to intracellular calcium regulation, APLNR[26], F2R[27], GPER1[19], JPH2[28], PKD2[29] and THY1/CD90[30] have already been investigated for their role in AAA. However, six additional genes are present in all pathways: ABL1, CALM1, CALM2, RYR2, SRI and CD19. Except for CALM2, all of them have been previously identified as DEGs in previous microarray studies between AAA and control samples[10–12,15], but none of these genes have been further investigated in either functional or epidemiological studies. These genes are closely related to intracellular calcium metabolism. ABL1 participates in both the release of stored intracellular calcium and extracellular calcium entry[31]. CALM1 and CALM2 code for two isoforms of the calmodulin protein, which plays a crucial role in the contraction of vascular and cardiac tissue through the detection of intracellular calcium[32]. RYR2 is mainly expressed in cardiac tissue and codes for the main regulator of sarcoplasmic calcium release[33]. SRI codes for the main binding protein of the RYR2 gene product[34]. The comparison between our AAA and control samples shows a downregulation of all these genes, suggesting a decrease in intracellular calcium levels in smooth muscle cells, consistent with loss of vascular contractility in the dilated aorta[35].

On the other hand, we observed upregulation of the CD19 gene in AAA. The activation of the surface protein encoded by CD19 triggers the release of intracellular calcium, which contrasts with the previous results[36]. However, CD19 is also a biomarker of B-cell development [36] which also play a key role in the development of AAA[37].

Among the pathways that were previously identified in differential expression analyses between AAA and control tissue is inflammation. There is a widely studied inflammatory component in AAA development involving both, adaptative and innate immune responses [17,18]. The presence of inflammatory infiltrates in AAA tissue have been widely demonstrated, which play a key role in the development of the disease[17,18]. The results of our cluster analysis also corroborated the association with the immune system after accounting for ischemic time, highlighting the key role of T cells on AAA development[38,39].

To better understand the effects of the inflammatory infiltrate in AAA development, we compared the proportions of inflammatory infiltrates between AAA and control samples. Some previous studies have analyzed the inflammatory infiltrates in AAA tissue[15,19,40]. In one study[15], immune cell proportions were estimated in AAA tissue layers (media and adventitia), without comparing with controls. Our results strongly corroborate their findings, suggesting that plasma B cells and M2 macrophages were the two most represented cell populations in both layers, and M1 macrophages, eosinophils, and activated dendritic cells were among the least represented cells in both layers. Surprisingly, resting mast cells emerged as our third most represented cell group, while in the layer-specific analysis, mast cells represented a small percentage of the total inflammatory cells. This contrasts with another study that compared whole-tissue samples between AAA and controls[19],where resting mast cells were among the most frequent cell groups in AAA samples. In addition, our results align with this study in detecting higher proportions of CD8 T-cells and lower proportions of resting NK cells in AAA samples. However, a discrepancy was noted in the levels of activated dendritic cells, which were more present in controls in our study but slightly higher in AAA samples in the previous work. Additional single-cell data demonstrated a higher proportion of T follicular helper cells and lower proportions of M1 and M2 macrophages in AAA samples compared to controls[40].

To rule out the effect of death as potential cause of variability, we used a reference work that evaluated the effect of death in blood samples,[41] which found higher levels of resting NK cells and CD8 T-cells in post-mortem samples. These results suggest that the increase in CD8 T-cells levels in AAA tissue could be even greater than the one we observed, and that the greater presence of NK resting cells in control samples could be due, in part, to their origin from organ donors. Consistent with this hypothesis, previous studies have shown that levels of NK cells are higher in the peripheral blood of AAA patients compared to controls,[42] and that these cells play a role in the development of the disease[18]. These results also suggest the existence of a highly cytotoxic environment led by CD8 T-cells in AAA tissue.

The lower levels of activated dendritic cells in the AAA samples compared to the controls was unexpected, given their established role as AAA inducers[17,19]. These results suggest that, although dendritic cells may participate in the development of AAA, they are not part of the inflammatory infiltrate.

### Study of alternative splicing between AAA and controls

This is the first study to elucidate the role of splicing in AAA development. We compared splice events between AAA and controls and identified eleven genes (SPP1, FHL1, GNAS, MORF4L2, HMGN1, ARL1, RNASE4, ASAH1, CYCS, HMGB1, SELENOP) with differentially represented splicing variants. We compared the splice events types identified in our comparison between AAA and controls samples with the presence of splicing events in the whole genome [21] and the observed proportions were comparable to the expected values genome-wide. The most frequent splicing types were alternative first exon (60 %) and skipping exon (20 %), while the least frequent were alternative 5’ splice-site (13.3 %) and mutually exclusive exon (6.67 %). On the other hand, it was surprising that our results did not include alternative last exon, alternative 3’ splice-site, and retained intron, despite their considerable genome-wide frequencies (10.72 %, 9.2 % and 3.54 %, respectively). This may be due to the limited sample size, as only 15 events were identified.

We observed seven genes that showed differential expression between AAA and control samples and had a splicing variant significantly more represented in AAA or controls (SPP1, FHL1, GNAS, MORF4L2, HMGN1, ARL1, RNASE4), indicating that splicing differences could be explaining the observed differential expression. Among these genes, SPP1 and FHL1 have been previously characterized in relation to AAA[43,44], whereas GNAS is a new DEG identified between AAA and control tissue. The evidence of differential splicing events validates GNAS as a new robust DEG between AAA and control tissue, and suggests that alternative splicing in this gene explains the differential expression and its implication to AAA. Finally, MORF4L2, HMGN1, ARL1 and RNASE4 although they have been identified in previous differential expression studies in relation to AAA[10–15], their specific role in AAA has not been studied. For these genes, our results contribute to understand the molecular mechanism leading to differential expression in AAA tissue.

SPP1 codes for the osteopontin protein, an important regulator of inflammation that has described functions in cardiovascular diseases[45]. SPP1 is more expressed in AAA tissue than controls, both in animal models and in humans, and it participates in AAA-associated extracellular matrix degradation[46,47] through the nuclear factor kappa B signaling pathway. It is also known that the SPP1 gene undergoes splicing and gives rise to 3 distinct isoforms osteopontin a, osteopontin b and osteopontin c, with specific characteristics, which have not been characterized in AAA. Consistent with previous data, our results found increased expression in AAA tissue, and identified for the first time that that skipping of exon 3 on SPP1 gene is more frequent in AAA than in controls, suggesting that this form of alternative splicing may be important for the development of AAA.

FHL1 codes for a protein that is highly expressed in skeletal and cardiac muscle. FHL1 has been shown to be a promising blood biomarker for human ascending thoracic aortic aneurysm as a modulator of metalloproteases.[44] Our findings, and those obtained in previous microarray studies[10–13,15], indicate that FHL1 levels are lower in AAA than in controls. We have detected for the first time that an alternative 5’ splicing-site form in this gene occurs more frequently in the control group, suggesting that AAA tissue would have reduced expression of this alternative isoform and reduced levels of FHL1, leading to higher risk of AAA development.

Although not previously found in transcriptomic studies, mutations in the *GNAS* gene have been studied in mice for their effect promoting AAA[48]. *GNAS* codes for the alpha subunit of the heterotrimeric G stimulatory protein (Gsα). Gsα may play a protective role in AAA development through regulation of vascular muscle tissue and is considered a potential therapeutic target[48]. Consistent with this protective role, our results confirmed lower expression *GNAS* levels in AAA. Moreover, our results add a mechanistic insight by revealing an alternative first exon splicing variant that occurs more frequently in controls and that could increase expression levels of the final protein and protect against AAA.

*MORF4L2* and *HMGN1* are DNA repair related genes, which had significantly lower and higher expression levels in AAA tissue compared to control tissue, respectively. *MORF4L2* has been associated with atheroma plaque progression in atherosclerosis[49]. Both genes present alternative splicing events that are less frequent in AAA tissue. Further work to elucidate the specific role of these genes in the risk of AAA development is warranted.

### Genes associated with AAA onset and progression

We identified eight genes (*EXTL3*, *ZFR*, *DUSP8*, *DISP1*, *USP33*, *VPS37C*, *ZNF784*, *RFX1*) that showed differential expression between AAA and control tissues, and also differential expression in AAA tissues of different diameters. Among these, *EXTL3*, *ZFR*, *DUSP8*, and *DISP1* have been previously identified as DEGs between AAA and controls but their role in AAA progression is novel.[10–15], and *USP33*, *VPS37C*, *ZNF784* and *RFX1* are novel DEGs. This could indicate that the genes may have a role beyond the disease onset and could be potential therapeutic targets to halt aneurysm expansion. Therefore, the potential contribution of these genes to the development of AAA needs to be thoroughly investigated.

Among the genes with higher expression at larger diameters, *USP33* encodes a deubiquitinating enzyme that has been associated with the development of hypertension[50], a known risk factor for AAA[51]. Proteins with deubiquitinating functions have been proposed as possible candidate genes for the treatment of AAA and other cardiovascular diseases[52]. Additionally, USP33 promotes the stabilization of beta2-adrenergic receptors, which promote vasodilation in smooth muscle[53]. We observed an increase in USP33 expression with increased AAA diameter, which is consistent with the functionality of beta2-adrenergic receptors. These results suggest that the USP33 gene may be a potential candidate for the treatment of AAA.

Among the genes that exhibit lower expression levels with larger diameters, we highlight DUSP8 and RFX1. DUSP8 is a phosphatase that negatively regulates the MAP kinase pathway, which is linked to cell differentiation and proliferation[54]. Previous studies have shown that DUSP8 expression levels are downregulated in mouse models of aortic dilatation[55], and the dual specificity phosphatase 8 protein (DUSP8) acts as a regulator of cardiac dynamics[54]. Our findings align with those obtained in the animal model of aortic dilatation, suggesting that DUSP8 may be a potential candidate gene for treating AAA by regulating both aortic dilatation and the immune system. RFX1 encodes a transcription factor that regulates genes involved in MHC class II[56]. In a previous bioinformatics study using microarray data, RFX1 was identified as a transcription factor that could potentially regulate DEGs between AAA and controls through its downregulation[57]. Previous research has shown that a decrease in RFX1 expression leads to activation of CD14^+^ monocytes in CAD patients[58]. Furthermore, CD14 protein plays a crucial role in recruiting macrophages during the early stages of AAA, and knockout mice for CD14 gene have been shown to resist the formation of AAA in two different models[59]. Finally, our expression results indicate that CD14 is upregulated in AAA patients compared to controls (FDR P-value = 0.002). In conclusion, these results suggest that low expression levels of the transcription factor RFX1 may lead increased CD14 expression, which plays a crucial role in the development and progression of AAA by recruiting macrophages.

Two previous studies have considered the diameter of AAA in differential expression analyses with aortic tissues. The first study[10] compared gene expression between small (n = 20) and large AAA (n = 29) with controls, but did not compare AAA of varying diameters. The second study[15] performed a correlation analysis on each gene between the diameter growth rate and gene expression in individuals (n = 24) with two aortic measurements, distinguishing between the media and adventitia aortic layers but did not identify any significant genes after multiple testing correction. The larger sample size in our study have enabled to identify for the first time genes that are associated to aneurysm progression, which could be potential therapeutic targets.

### Identification of haplotypes associated with AAA risk

We performed a clustering analysis of enriched biological pathways with the 1,815 genes with significant differences in the allele specific expression patterns between AAA and controls. The cluster analysis revealed a strong association with immune system pathways, particularly those associated with T cells. These results are consistent with the clustering analysis of DEGs between AAA and controls and demonstrate how specific haplotypes determine the differential expression of genes associated with AAA in the diseased tissue. By combining allele specific expression information on AAA samples with the allelic specific expression results between AAA and controls, along with information from a previously published GWAS[7], we explored the associations between genetic variants associated with AAA risk and gene expression. This approach aimed to unravel the association between genetic haplotypes and gene expression as determinants of AAA risk.

SNURF is the only gene that showed significant allelic specific expression in more than five of the twelve studied individuals with AAA and differential allelic specific expression patterns between AAA and controls suggesting the existence of a specific haplotype associated with less expression in AAA and leading to higher risk of AAA. SNURF codes for an open reading frame of the SNRPN gene, with which it forms a bicistronic gene[60]. Both genes exhibit significantly lower expression in AAA than in controls in our data. Moreover, we observed that rs705 single nucleotide polymorphism (SNP) (minor allele frequency (MAF) (C) = 0.45) determined SNURF expression in eight out of twelve AAA samples, where the T allele was associated with double expression amounts compared to the C allele. Additionally, rs705 is an eQTL in blood of the SNRPN gene. Although causality cannot be derived from these results, these results suggest a possible effect of the haplotype containing the rs705 T allele on this locus on SNURF expression, resulting in higher risk for AAA development.

Among the genes with significant allele specific expression in the twelve analyzed AAA samples that did not exhibit significant allele specific expression patterns between AAA and control groups in our analyses, we selected those that were part of a locus associated to AAA in the most recent GWAS on AAA risk[7]. THBS2 encodes for thrombospondin 2, a protein that regulates cell-cell and cell-extracellular matrix interactions and has been studied in relation to multiple cardiovascular diseases[61]. THBS1, a member of the same family, has been identified as a regulator of AAA in animal models[62]. Our allele specific expression results suggest that the expression of this gene could be associated with AAA development, supporting the implication of the gene in AAA risk. Two common genetic variants (rs58023137 (MAF (T) = 0.22) and rs9505895 (MAF (A) = 0.2)) were identified in previous GWAS studies[7] and in our allele specific expression results. For both variants, the GWAS risk allele was associated with higher expression gene levels in one individual with AAA. Additionally, these two variants were also eQTLs in aortic tissue regulating THBS2 gene expression, confirming the known association between thrombospondin and AAA[7], and demonstrating the presence of risk haplotypes associated to increased expression and risk of AAA.

Regarding the SPP1 gene, we found six genetic variants (rs35893069 (MAF (T) = 0.1), rs6839524 (MAF (G) = 0.12), rs4754 (MAF (C) = 0.28), rs1126616 (MAF (T) = 0.27), rs1126772 (MAF (G) = 0.21), rs9138 (MAF (C) = 0.22)) with allele specific expression among the twelve AAA samples. As previously discussed, the SPP1 has been associated to AAA risk due to the role of the protein osteopontin, which it encodes, in the degradation of the extracellular matrix that is characteristic of AAA[46,47]. However, none of the six genetic variants were found to be significant in the previous GWAS[7], and therefore no association with allelic expression could be established.

### Strengths and limitations and comparison with previous studies

This is one of the largest studies of differential expression between AAA tissue samples and controls. However, due to the difficulty in obtaining aortic tissue samples, the sample size is still limited compared to transcriptomic studies in more accessible tissues, which may have limited our power. Previous studies comparing transcriptomics between AAA and control samples have been limited to the use of expression microarrays[10–15]. In this study, we used RNAseq to obtain transcriptomic information. RNAseq provides greater sensitivity, a wider range of detection of both high and low expression genes, and is not limited to microarray probes[9,63], which allowed us to detect more DEGs. Additionally, RNAseq technology allowed for a more precise study of alternative splicing compared to microarrays.

Accounting for ischemic time enabled us to more accurately identify biological processes associated to AAA, thereby minimizing differences in sample origins between AAA and controls. At the same time, despite ensuring more robust findings, the elimination of these genes could have been too rigorous and imply the loss of some biological pathways associated with AAA that, at the same time were affected by ischemic time.

The significant phenotypic differences in age and sex between individuals recruited as AAA and controls are a limitation of the study. Moreover, the analyses of the allele specific expression are limited by the small sample size (n = 12) and the requirement for the individuals to be heterozygous in order to study the genetic variants. Therefore, further validation is necessary.

However, this is the first study analyzing allele specific expression in AAA and our results emphasize the value of this new dataset combining genetic and expression data for the study of AAA.

Finally, the conclusions presented in this article are based on the results obtained from a cohort of European-ancestry individuals only. Further analyses in different ancestries will determine the generalization of these results.

## CONCLUSIONS

Our analysis of the whole transcriptome has enabled us to identify numerous novel genes that were not previously detected in microarray studies. Additionally, our efforts to account for the impact of ischemic time have provided more robust implicated biological pathways that lead to AAA development. Among these pathways, ATP synthesis regulation (i.e. genes encoding subunits I, III and IV of the electron transport chain), has been associated for the first time in transcriptomic studies of AAA tissue, although this pathway was previously studied in relation to AAA in previous studies[23,24]. Additionally, the study of differential splicing processes between AAA and controls have revealed novel molecular processes involving already known genes in relation to AAA, such as SPP1, FHL1 or GNAS.

The study also analyzed the differential expression of genes in individuals with AAA of different diameters, providing valuable insights into the underlying molecular mechanisms contributing to AAA progression. Further research on these genes may lead to potential treatments against aneurysm expansion, which increases the risk of rupture. Finally, the analysis of differential allele specific expression in twelve AAA has allowed the identification of haplotypes associated with expression of certain genes and AAA risk, providing evidence for their involvement in disease, and shedding light into the molecular mechanism.

Overall, this study provides a comprehensive exploration of AAA expression patterns, revealing key insights into the pathophysiology of AAA initiation and progression. RNAseq was used to conduct one of the largest differential expression analyses to date, uncovering numerous genes associated with AAA. Our consideration of ischemic time and AAA diameter improved the precision in identifying biological processes associated with AAA onset and progression. Furthermore, our analysis of differential allele specific expression has identified genetic haplotypes that influence gene expression on AAA tissue, which advances our understanding of AAA genetic background. These findings contribute to future research and potential advances in precision medicine to reduce AAA progression and mortality risk.

## METHODS

### Subjects

We used a total of 140 human abdominal aortic samples from 96 patients diagnosed with infrarenal AAA and undergoing open surgery for AAA repair at Hospital de la Santa Creu i Sant Pau and Hospital del Mar (Barcelona) and 44 controls. The study participants were obtained from TABS cohort, which includes genomic, transcriptomic, clinical, and maximum aneurysm size data, from AAA patients and healthy individuals. Maximum aortic diameters were obtained from computed tomography or ultrasound images. Genomic data were obtained through genotyping using the Infinium Global Screening Array-24 v2.0 from Illumina (San Diego, California) (coverage 665,608 variants) and imputation to the TOPMED Reference Panel. Only variants with imputation quality > 0.3 were used for the allele specific expression analyses. Healthy abdominal aortas were obtained from 21 male and 23 female multiorgan donors (Table 1).

### Sample processing

A portion of tissue sample was placed in RNAlater solution (Qiagen GmbH, Hilden, Germany) and stored for 24 hours at 4 °C before long-term storage at −80 °C until further processing. For RNA isolation, tissues were then homogenized in 1 ml Trizol (ambion, Carlsbad, CA) in the FastPrep-24 homogenizer and Lysing Matrix D tubes (MP Biomedicals, Solon, OH) and RNA was purified using PureLink RNA Mini Kit (Invitrogen, Carlsbarg) following the manufacturer’s recommendations. RNA concentration was measured using Nanodrop 200 (Thermo Scientific).

RNA integrity was assessed on Agilent 2100 Bioanalyzer (Agilent, Santa Clara, CA, USA). RNA integrity number (RIN) was recorded and only samples with a RIN higher than 6 were used.

### RNA sequencing

We performed sequencing analyses using Illumina NovaSeq 6000, with a read length of 150 bp and paired-end sequencing. Two sequencing runs were performed to reduce the variability of the technical variables. First, AAA and control samples were randomized between sequencing plates. Second, only AAA were sequenced with as little technical variability as possible. In all sequencing runs, a minimum of 30 million reads were required, repeating the sequencing on a new plate if this limit was not reached.

### Alignment, quantification, and quality control

We used STAR v.2.5.3a[64] to perform the alignment on the reference genome version GRCh38 and we then used RSEM v1.3.0[65] for gene quantification. For both analyses, we used gene models from GENCODE v26 gene annotation file[66]. In total, 58,219 genes were quantified. Of these, we selected protein coding genes and long non-coding RNA (lncRNA) genes that had been either experimentally validated (level 1 annotation) or manually annotated (level 2 annotation)[66], resulting in 27,290 genes (19,777 protein-coding and 7,513 lncRNA) for further analyses. Gene quantifications were expressed as Transcripts Per Million (TPMs), which were obtained by normalizing for gene length first, and then for sequencing depth. This ensured that the sum of all TPMs in each sample was the same, facilitating the comparison between samples. All samples reached a minimum of 10 million reads aligned to the reference genome with STAR (Additional file 1: Figure S5A-5B)[64].

As part of the quality control, we also checked that the reported sex of the samples matched the biological sex of the sequenced data. To do this, we compared the expression levels of the XIST gene, which regulates the X chromosome inactivation mechanism in females and has null expression in males, with the expression of male exclusive genes, calculating an average expression of Y chromosome genes. One sex mismatch sample was eliminated from the study (Additional file 1: Figure S5C).

### Differential expression analysis

Before conducting differential expression analyses, we normalized the TPMs counts using quantile normalization and removed lowly expressed genes by removing genes with less than 0.5 TPMs in more than 50% of the samples. For the comparison of AAA against control samples, we kept 14,675 genes and removed 12,615 genes. For the comparisons among AAA using the AAA-only sequencing panel, we kept 14,779 genes and removed 12,511 genes.

To evaluate the impact of technical covariates on the results, we performed a Principal Components (PC) Analysis on all samples and tested the correlation between the PC Analysis and all the technical covariates. Technical covariates that had significant correlations (p-value < 0.05) with the first four PC were included in the analyses as fixed effect covariates. Additionally, all comparisons were adjusted for age and sex. To preserve the regulatory effects that act through smoking, we did not include smoking as a covariate (Additional file 10: Table S9).

First, we calculated DEGs between AAA and controls using a linear regression, including age, sex, flow cell type, flow cell lane, GC mean content (GC mean), RIN, percentage of RNA fragments > 200 (DV200), and Qubit as covariates. Date of creation of the library was not included due to its high correlation with the case/control variable (Pearson’s correlation = - 0.74) (Additional file 1: Figure S6A-6B)). Second, we calculated DEGs between aneurysms of varying diameter using a linear regression, including age, sex, date of creation of the library, batch number, GC mean, RIN, DV200 and Qubit. We corrected for multiple testing in both analyses using Benjamini-Hochberg false discovery rate (FDR) method and considered significant DEGs those with an adjusted p-value below 0.05[67].

We then performed a linear regression model to identify genes whose expression could be altered by ischemic time, using ischemic time information on artery aorta tissue samples from GTEx V8 data[16]. We corrected the linear regression model for age, sex, RIN, type 2 diabetes, body mass index, autolysis score, center, sequencing protocol, sequencing platform, and genotyping PC. To determine the genes affected by ischemic time, we corrected for multiple testing and selected genes with an FDR-adjusted p-value lower than 0.05[67]. All analyses were performed in R.

### Enrichment analysis

After identifying DEGs between AAA and control samples and between AAA of varying diameters, we performed enrichment analyses using the R package ‘clusterprofiler’[68,69] on the Gene Ontology (GO) databases for Biological Process, Cellular Component and Molecular Function, as well as the Kyoto Encyclopedia of Genes and Genomes (KEGG)[70]. We corrected for multiple testing using the Benjamini-Hochberg FDR method and identified significantly enriched pathways with an adjusted p-value below 0.05[71]. We used ‘aPEAR’[72] to perform a cluster analysis of redundant pathways with a minimum cluster of size of 15 and hierarchical clustering. The enriched pathways and genes were visualized using ‘enrichplot’[73].

### Study of the inflammatory infiltrate

We investigated the differences in the proportions of inflammatory infiltrates between AAA and control samples using CIBERSORTx[74]. CIBERSORTx compares RNAseq data with a reference expression database of selected cell types, to estimate the proportion of each cell type. The residuals of our RNA-seq data were calculated using a linear regression that included all covariates except the status variable. We used the residuals and the ‘lm22’ signature matrix, that contains expression data of 547 genes in 22 inflammatory cell types from microarray studies, and can be used to distinguish inflammatory cell populations in RNA-seq data[74]. We conducted a t-test to compare the proportions of each cell type between AAA and controls. A p-value threshold corrected by multiple testing using Bonferroni for the number of cell types was set at p-value < 2.27 x 10^-3^ (0.05 / 22).

### Alternative splicing

We used the SUPPA2 software to identify differences in splice events between AAA and control tissue samples. SUPPA2 can identify seven different splice events including skipping exons, mutually exclusive exons, alternative 5’ or 3’ splice sites, retained introns, and alternative first or last exons. Based on gene annotation from GENCODE v26,[66] we computed the proportion of splice inclusion for the TPMs counts in each splice event by dividing the number of TPMs of one form of the event by the total number of TPMs. Finally, the magnitude of splicing change was calculated by subtracting the proportion of splice inclusions between AAA and controls. Significant alternative splicing events were selected based on a magnitude of splicing change higher than 0.1 and FDR-corrected p-values lower than 0.05, using the default parameters for calculation.

### Allelic specific expression

Allele specific expression was investigated using PHASER[75] on twelve AAA samples with genotype data. Allele specific expression consists of the analysis of the differences in the expression levels of the different haplotypes present in a heterozygous individual. We quantified allele specific expression at the gene level using the GENCODE V26 gene annotation[66]. To reduce the effect of the known mapping bias towards the reference allele[76], we performed an additional STAR mapping step with WASP filtering[77].

We used the allele specific expression data obtained by the GTEx consortium to compare allele specific expression between our AAA samples and GTEx control tissues[78]. To do this, we compared the proportional expression of each allele between our AAA samples and the GTEx controls, using a non-parametric Wilcoxon test[79]. Then, to identify genes with distinct allele specific expression patterns, we corrected for multiple comparisons using FDR[67]. We performed enrichment and cluster analyses in all genes showing FDR adjusted p-values lower than 0.05.

## ABBREVIATIONS

AAA: abdominal aortic aneurysm
DEGs: differentially expressed genes
TABS: triple A Barcelona study
LncRNA: long non-coding
RNA TPMs: transcripts per million
PC: principal component
GC mean: GC mean content
DV200: percentage of RNA fragments > 200
FDR: false discovery rate
GO: gene ontology
KEGG: Kyoto encyclopedia of genes
MHC: major histocompatibility complex
GWAS: genome-wide association study
ATP: adenosine triphosphate
Gsα: heterotrimeric G stimulatory protein
DUSP8: dual specificity phosphatase 8 protein
SNP: single nucleotide polymorphism
MAF: minor allele frequency

## DECLARATIONS

### Ethics approval and consent to participate

The study was approved by the Hospital de la Santa Creu i Sant Pau Ethics Committee (IIBSP-OMI-2019-102). All patients gave written informed consent prior to surgery to participate in the study. The study conformed to the principles of the Declaration of Helsinki.

### Consent for publication

Not applicable.

### Availability of data and materials

The personal data used for this study is available from the corresponding author on reasonable request for collaborations provided it complies with the ethical permits of the study. All other data supporting the findings of this study is available within the paper and its Supplementary Information. Code used for data preparation and analysis is available at https://github.com/Gerardts9/RNAseq.

### Competing interests

The authors declare that they have no competing interests.

### Funding

This work was supported by a grant from the Spanish Ministry of Science and Innovation (PID2019-109844RB-I00). The genotyping service was carried out at the Genotyping Unit-CEGEN in the Spanish National Cancer Research Centre (CNIO), supported by Instituto de Salud Carlos III (ISCIII), Ministerio de Ciencia e Innovación. CEGEN is part of the initiative IMPaCT-GENóMICA (IMP/00009) cofunded by ISCIII and the European Regional Development Fund (ERDF). GT-S is supported by the Pla Estratègic de Recerca i Innovació en Salut (PERIS) grant from the Catalan Department of Health for junior research personnel (SLT017/20/000100). MS-L is supported by a Miguel Servet contract from the ISCIII Spanish Health Institute (CPII22/00007) and co-financed by the European Social Fund. DD was funded by a Tenovus Scotland Research PhD studentship, T19-06.

### Author’s contributions

AV, MC, and MS-L conceived and designed the study. BS, JD, OP, LC, LN and J-RE generated the clinical database and supplied the samples. GT-S and DD performed the analyses. GT-S, AB, AV, MC, and MS-L interpreted the results. GT-S, AV, MC, and MS-L wrote the manuscript. All authors read and approved the final manuscript.

## Acknowledgments

We acknowledge the AAA patients at Hospital de la Santa Creu i Sant Pau and Hospital del Mar who participated in this study, and the multiorgan donor families who made this research possible through their generous consent.

## Author’s information

AV, MC, and MS-L contributed equally to this work as senior authors.

## SUPPLEMENTARY INFORMATION

**Additional file 1:** Figures S1-S6 Contains Supplementary Figures S1-S6.

**Additional file 2:** Table S1

Complete differential expression results between AAA and controls.

**Additional file 3:** Table S2

GO functional enrichment analysis results between AAA and controls (including genes affected by ischemic time).

**Additional file 4:** Table S3

KEGG functional enrichment analysis results between AAA and controls (including genes affected by ischemic time).

**Additional file 5:** Table S4

Differential expressed genes by ischemic time in GTEx samples.

**Additional file 6:** Table S5

GO functional enrichment analysis results between AAA and controls (excluding genes affected by ischemic time).

**Additional file 7:** Table S6

KEGG functional enrichment analysis results between AAA and controls (excluding genes affected by ischemic time).

**Additional file 8:** Table S7

Results of the alternative splicing study between AAA and controls.

**Additional file 9:** Table S8

Complete differential expression results between AAA of different diameter.

**Additional file 10:** Table S9

Correlation analysis between biological and technical covariates.

## Notes

### Competing Interest Statement

The authors have declared no competing interest.

