## Supplementary Figures for "Differential expression analyses on aortic tissue reveal novel genes and pathways associated with abdominal aortic aneurysm onset and progression"

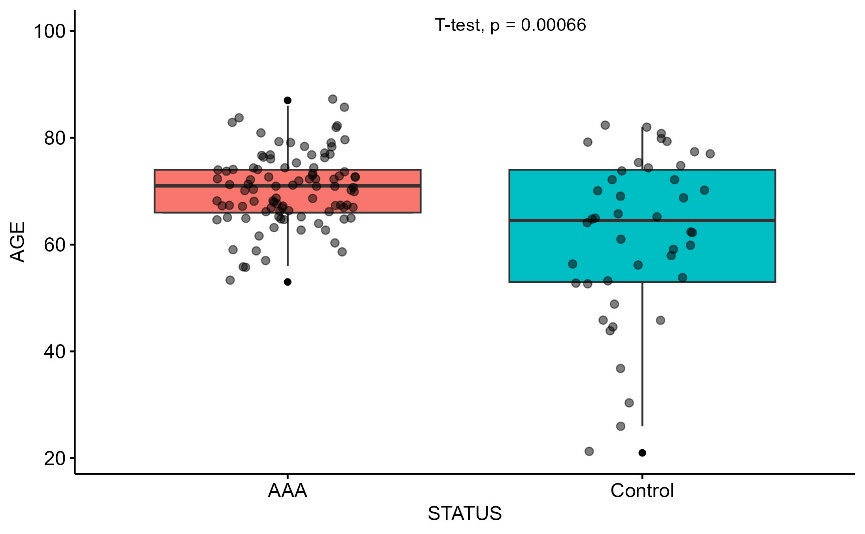

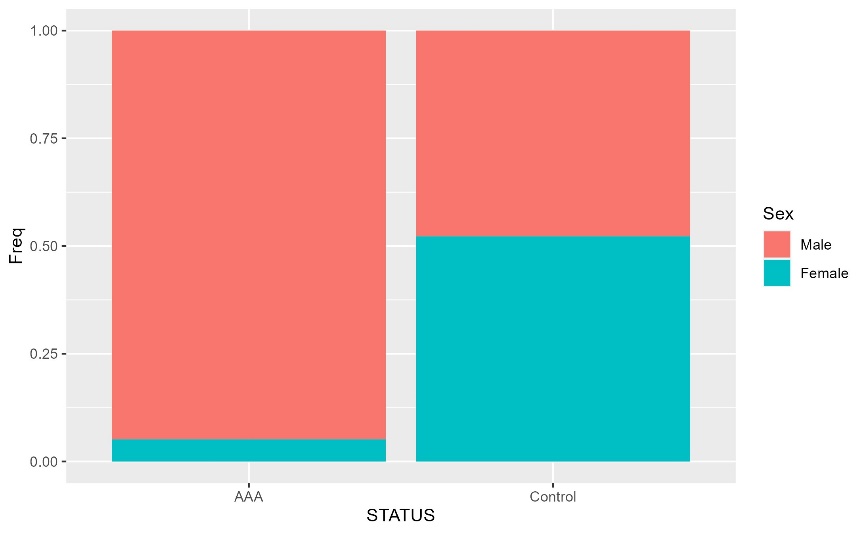

B

A

C

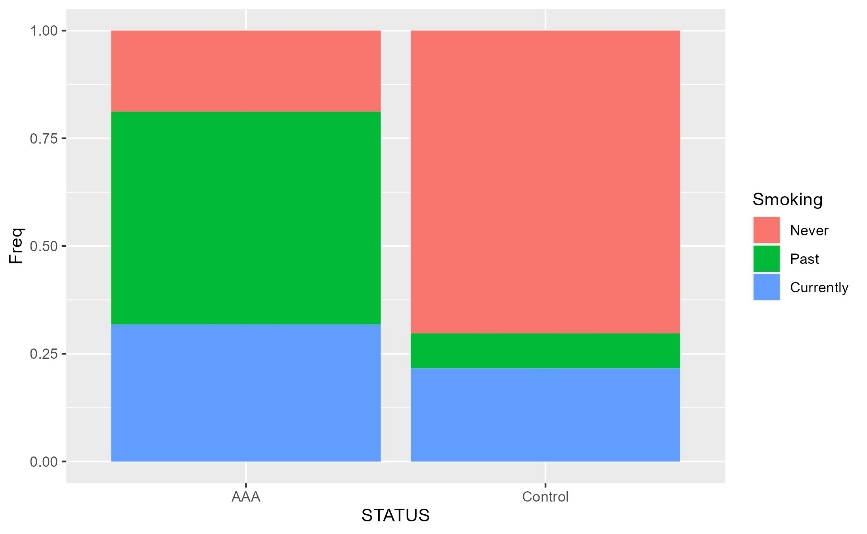

**Supplementary Figure S1: Individuals statistical description plots.** A) Barplots comparing the proportion of males and females between AAA and controls B) Boxplots comparing the age between AAA and controls C) Barplots comparing the smoking status between AAA and controls

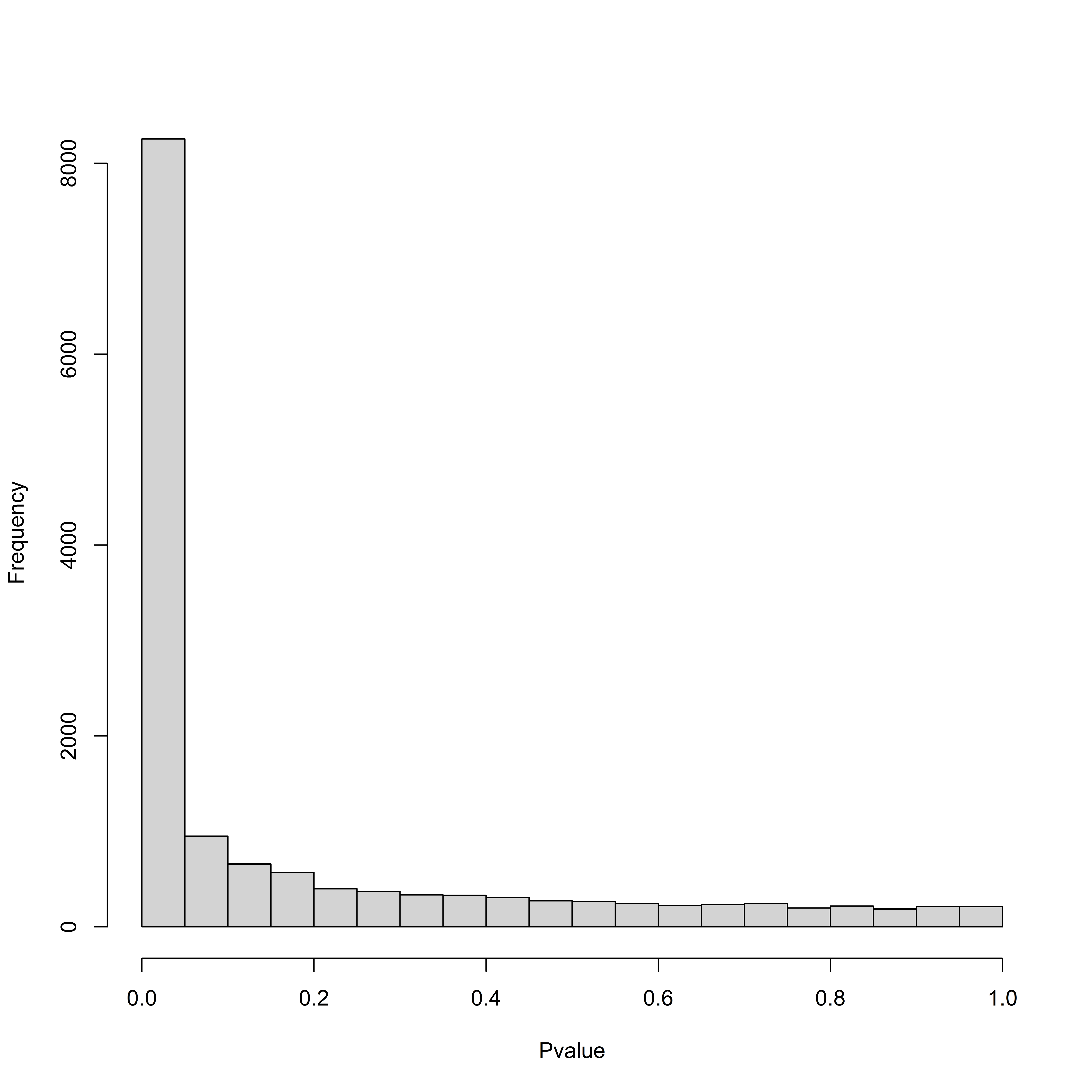

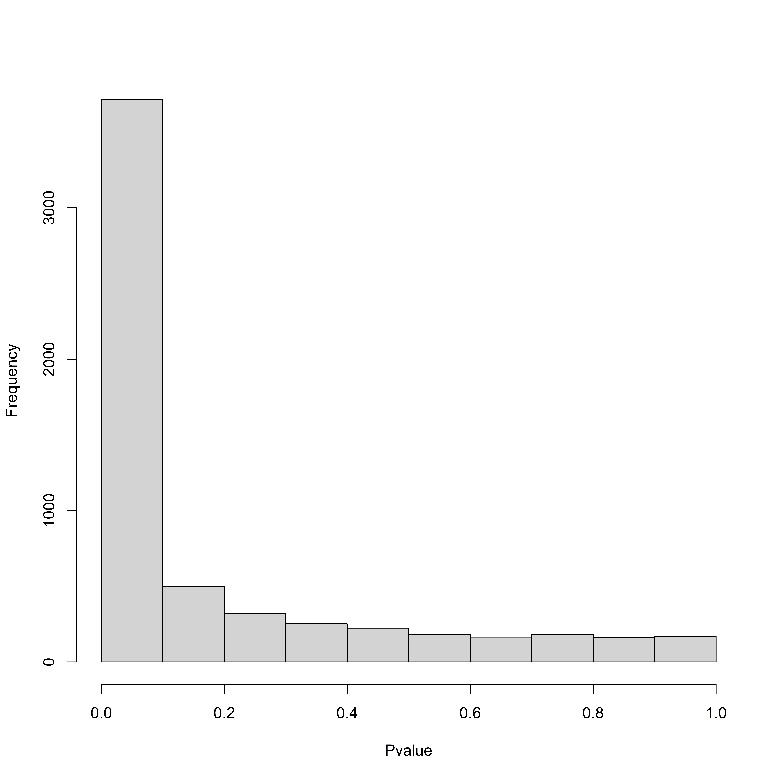

B

A

Pi1 = 0.72

Pi1 = 0.72

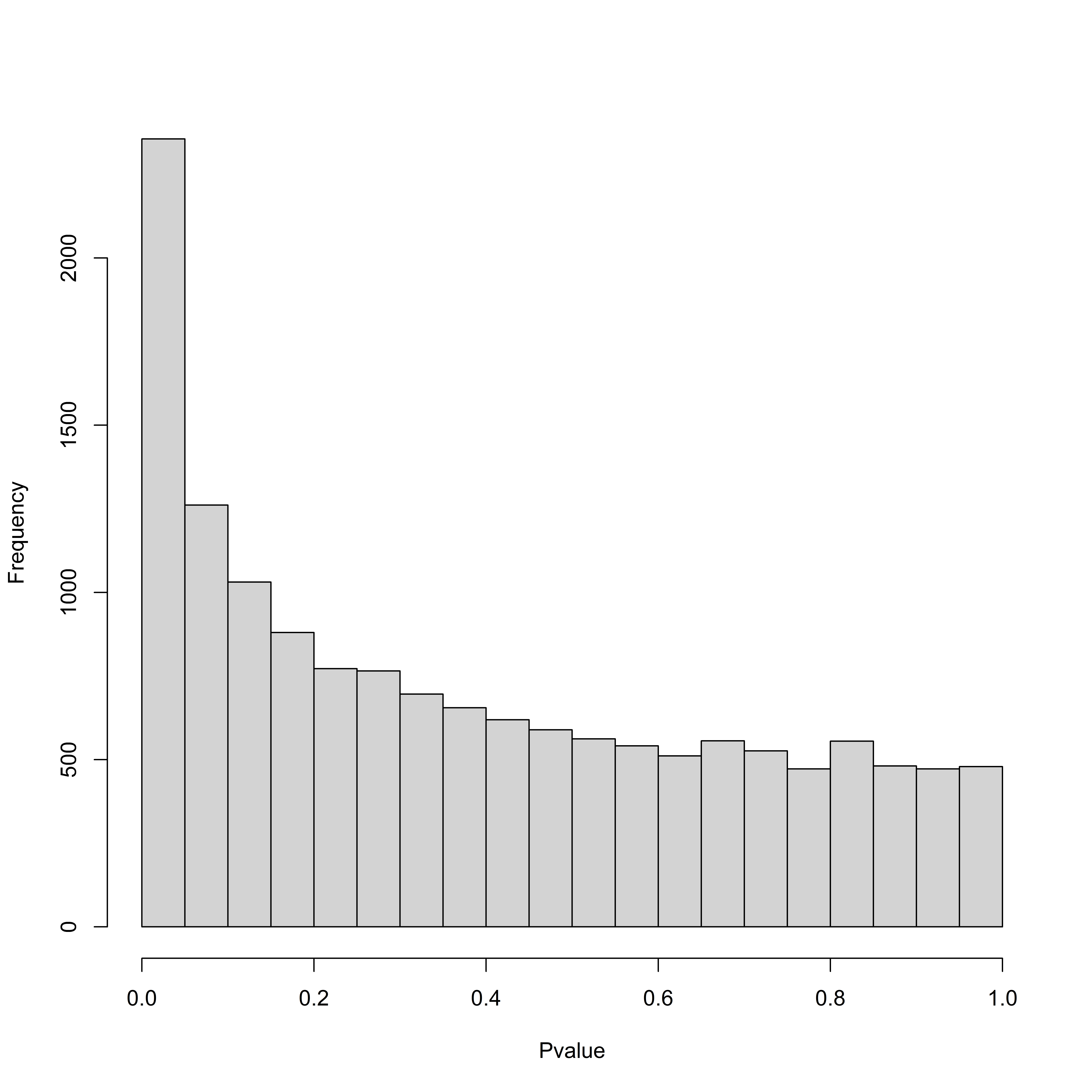

C

Pi1 = 0.36

**Supplementary Figure S2: Differential expression p-values histograms.** Distribution of p-values obtained in the linear regressions performed to identify differentially expressed genes in A) AAA against controls B) AAA against controls, after removing genes affected by ischemic time C) aneurysms of varying diameter. Pi1 values are an estimation of the true non-null p-values considering the whole distribution of p-values obtained with the ‘qvalue’ R package.

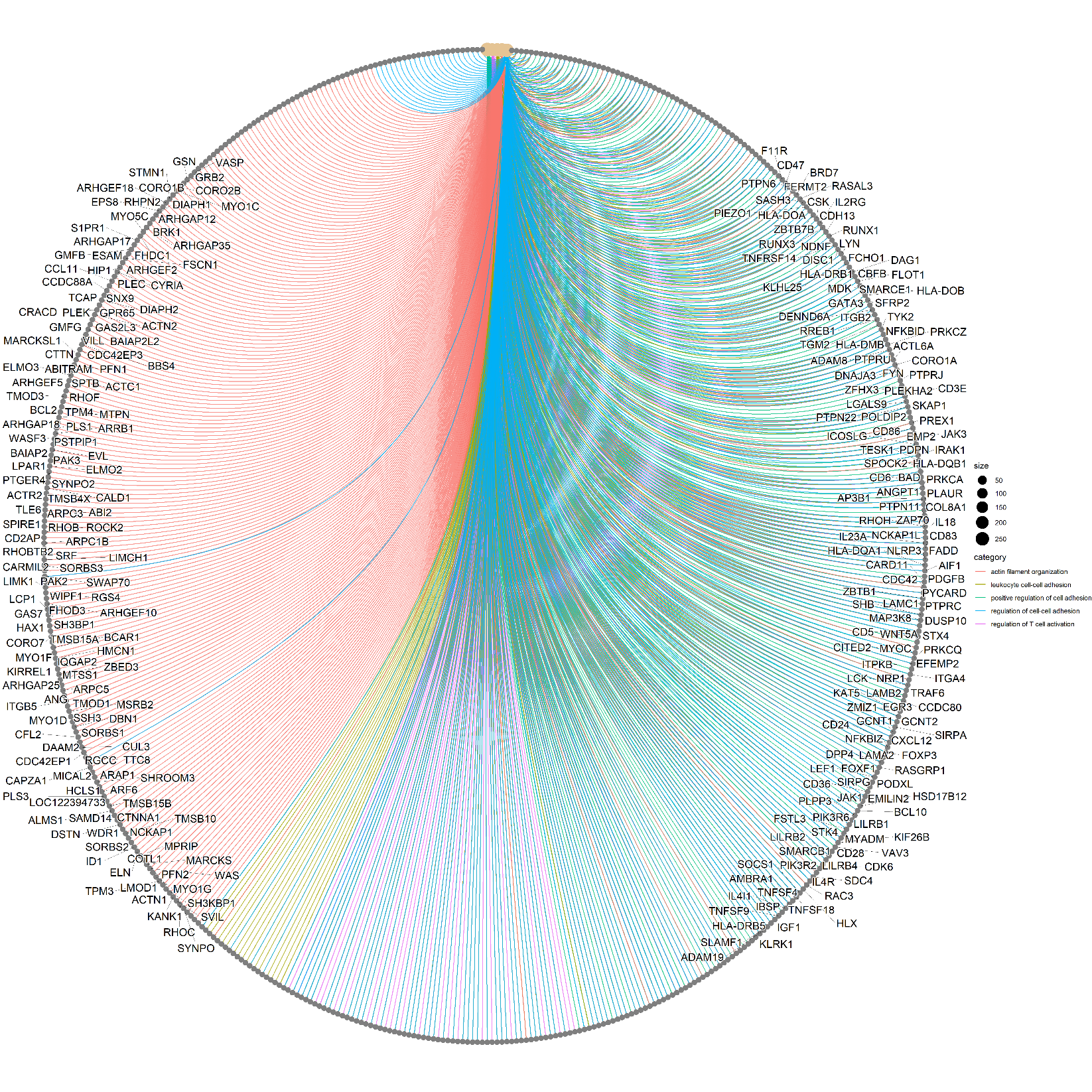

A

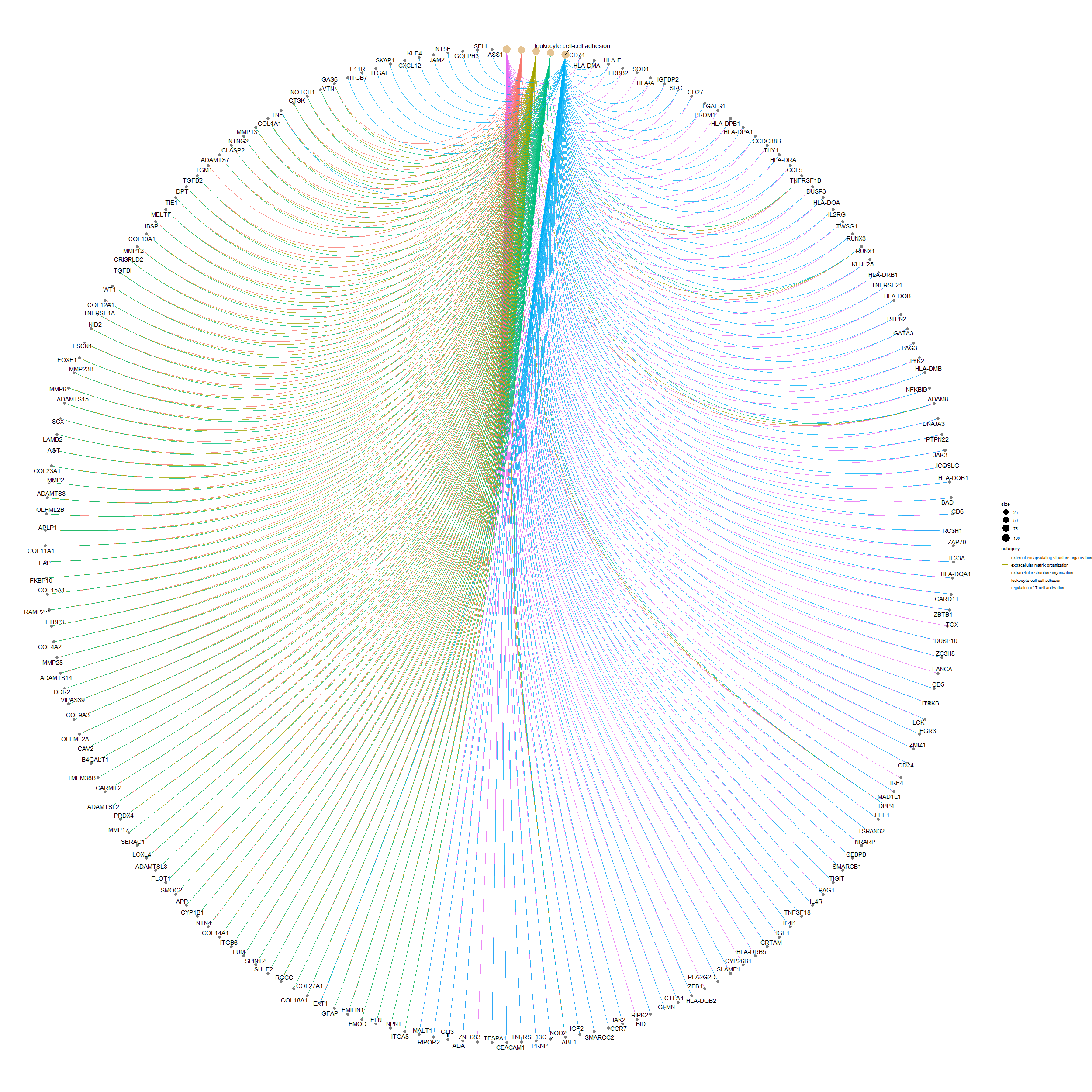

B

**Supplementary Figure S3: Network plot depicting the linkage of genes and biological pathways from gene ontology terms.** Graphical representation of the enriched pathways and genes obtained in the gene ontology analysis performed with the R package 'clusterpofiler' and plotted with 'enrichplot'. A) Using all differentially expressed genes between abdominal aortic aneurysm patients and controls B) Using differentially expressed genes between abdominal aortic aneurysm patients and controls after excluding genes affected by ischemic time.

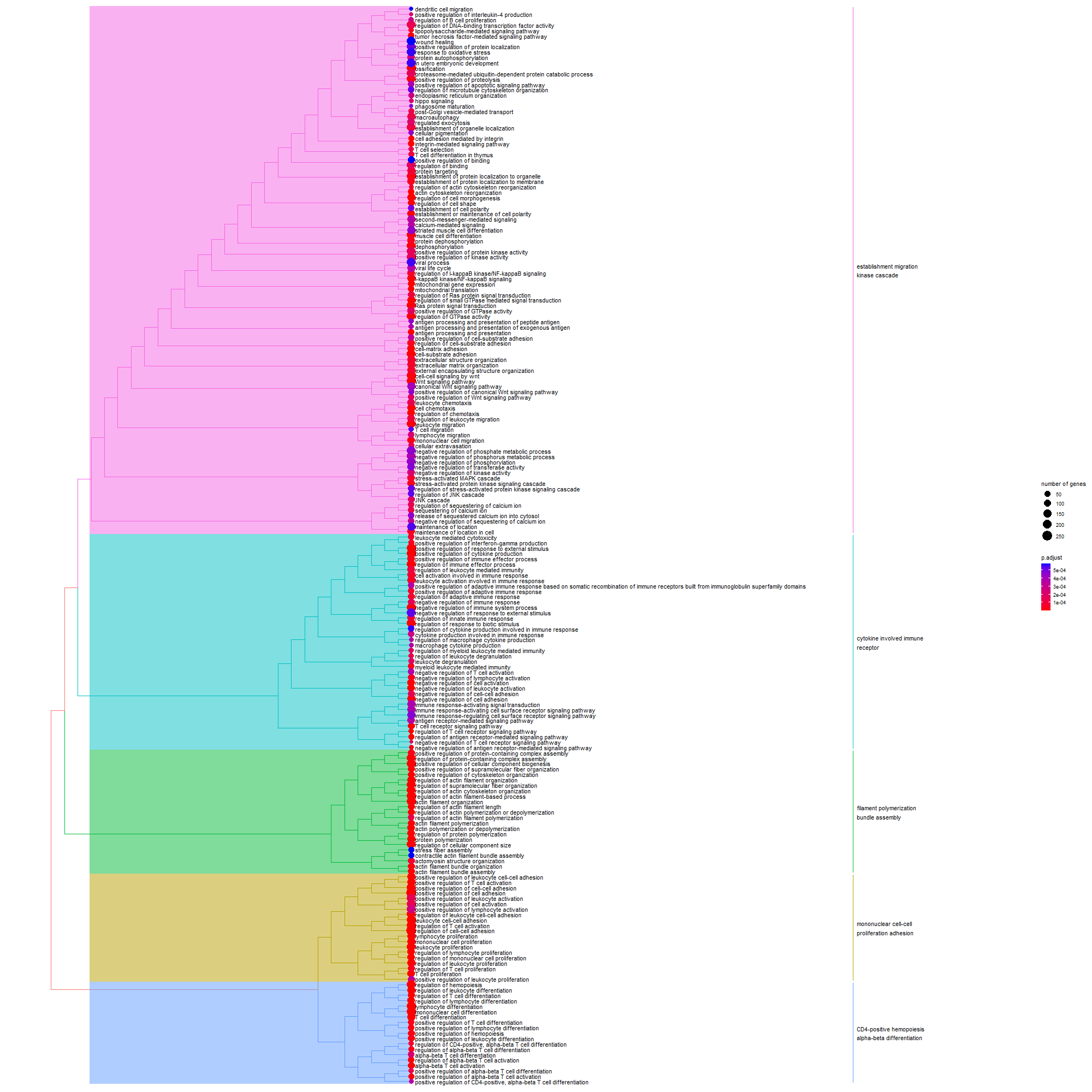

A

B

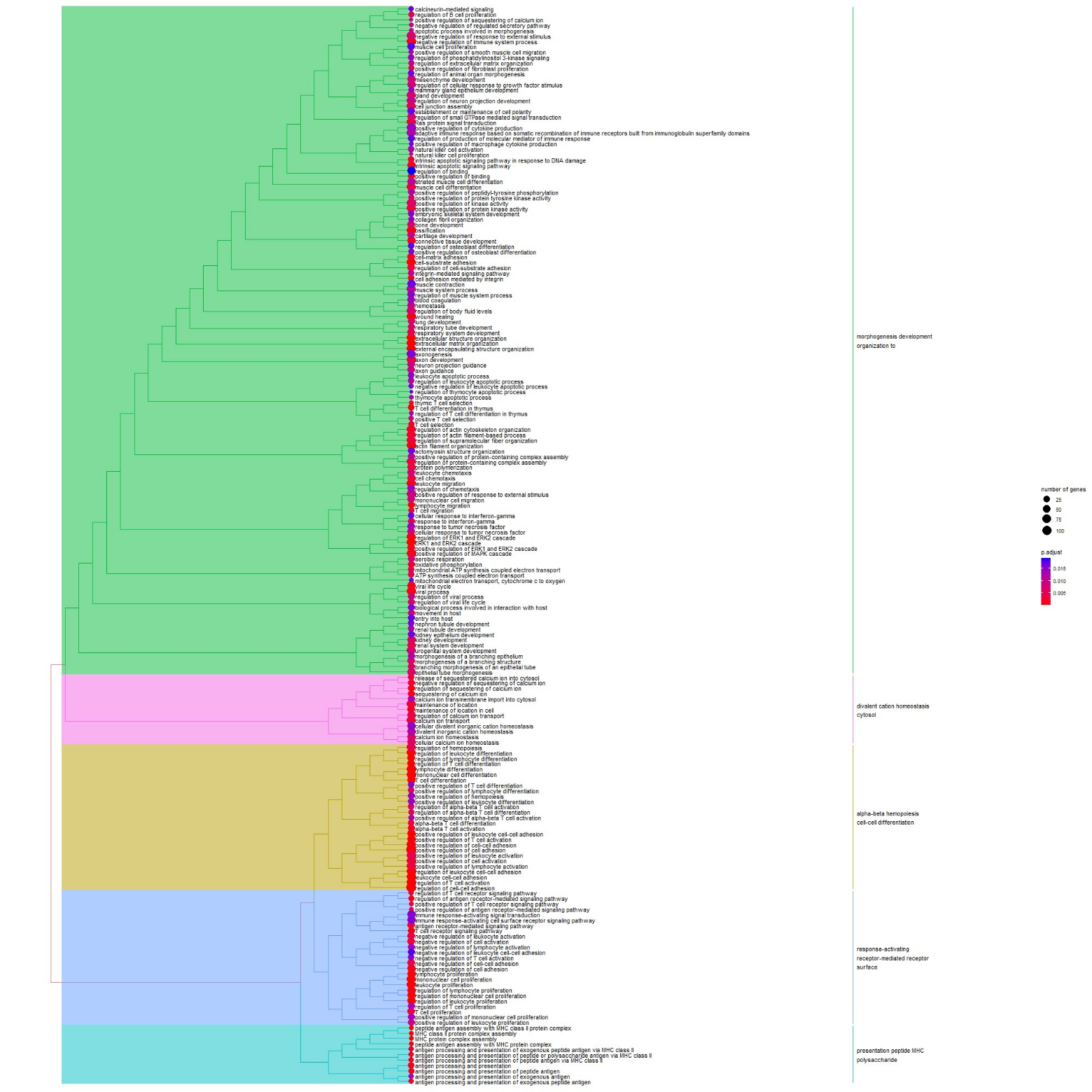

**Supplemental Figure S4: Tree plot of the hierarchical clustering of enriched GO terms.** Graphical representation of the enriched pathways obtained in the gene ontology analysis performed with the R package 'clusterpofiler' and plotted with 'enrichplot'. A) Using all differentially expressed genes between abdominal aortic aneurysm patients and controls B) Using differentially expressed genes between abdominal aortic aneurysm patients and controls after excluding genes affected by ischemic time.

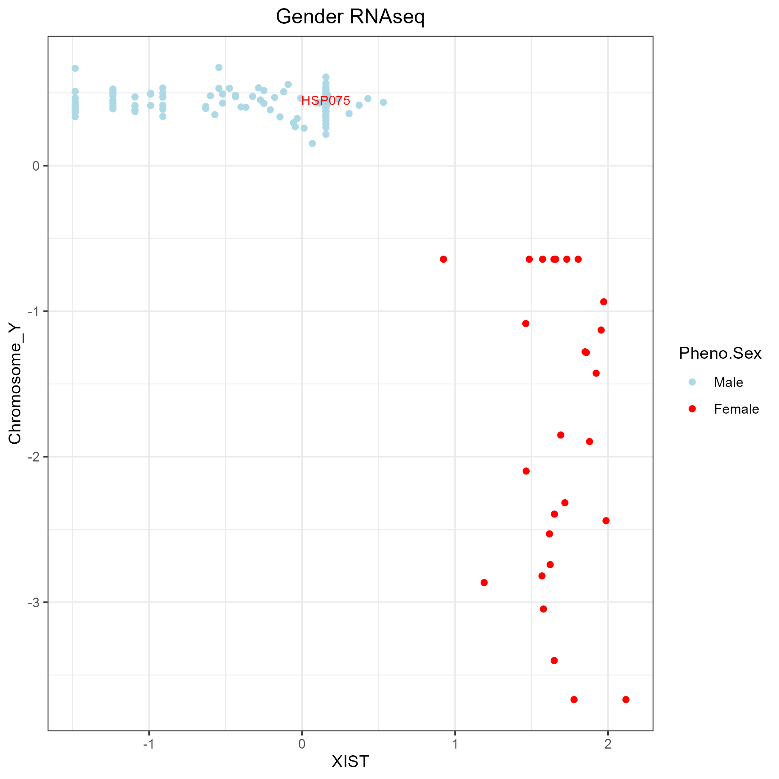

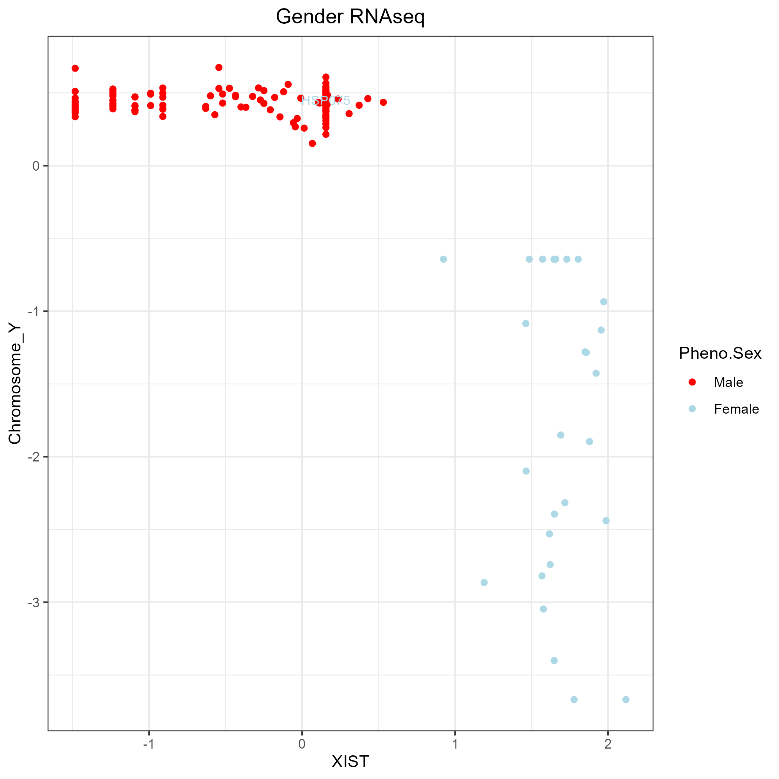

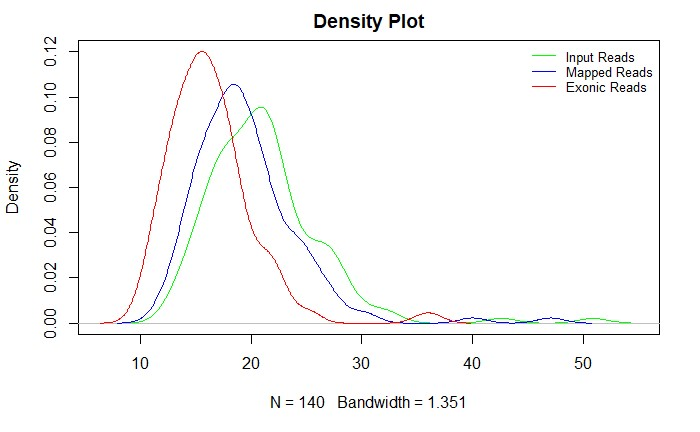

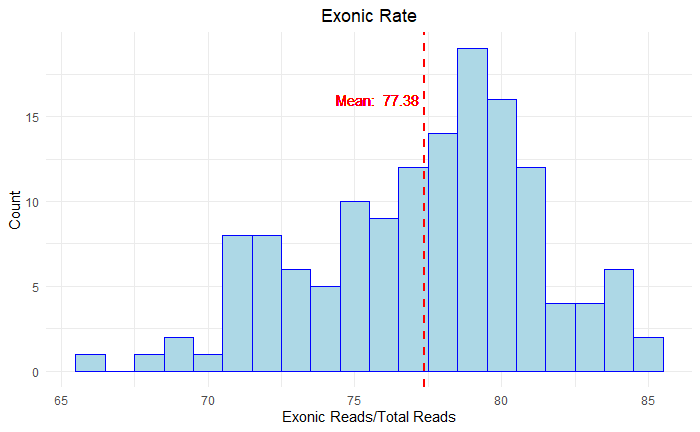

C

A

B

**Supplementary Figure S5: Quality control plots.** A) Density plot representing the number of input reads (green), mapped reads (blue) and exonic reads (red) obtained after mapping with the reference genome B) Histogram of the exonic rate, calculated using the formula Exonic Reads / Total Reads. The vertical red line indicates the mean value of the exonic rate for all samples. C) Comparison of *XIST* gene expression, which has null expression in males, with the mean expression of genes on the Y chromosome to detect sex mismatches. HSP075 was detected as a sex mismatch and removed from the study.

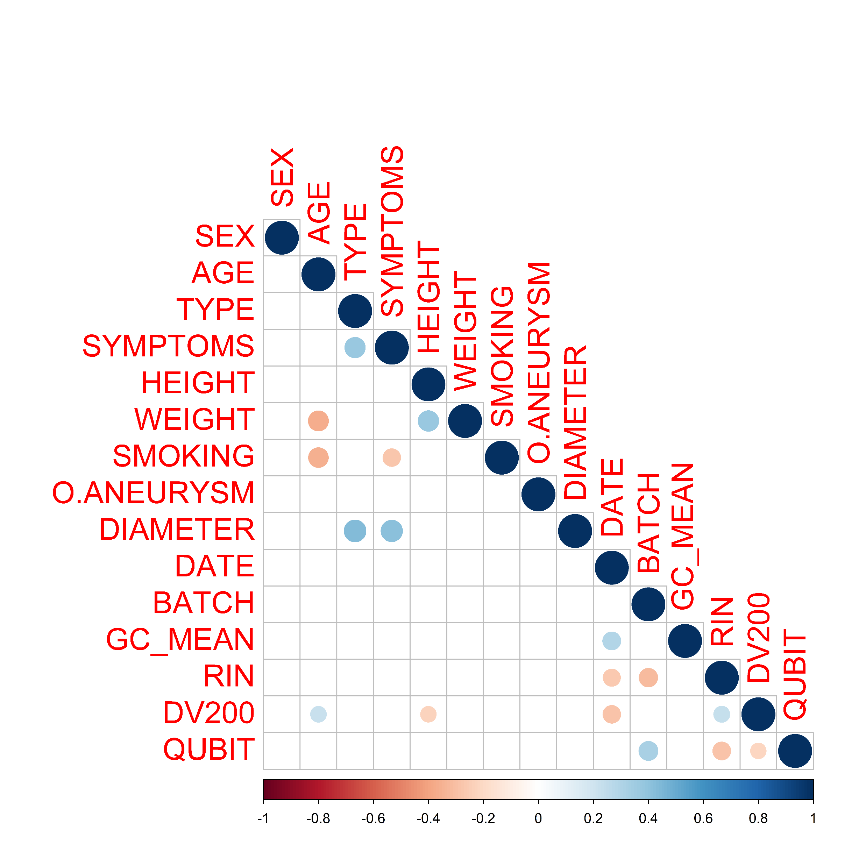

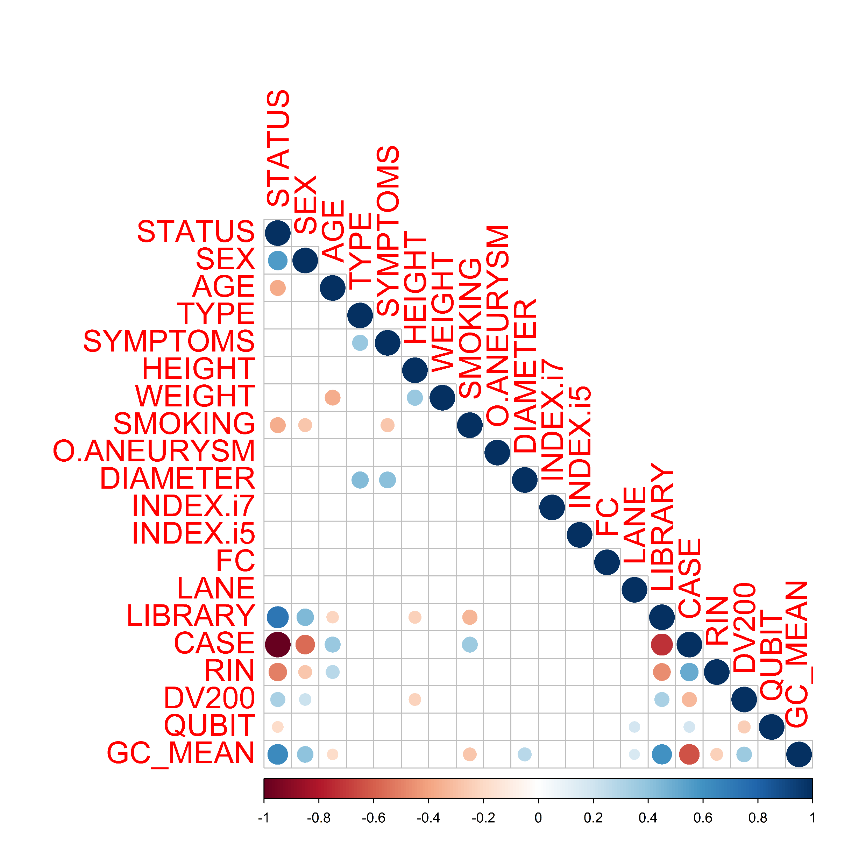

**Supplementary Figure S6: Pearson’s correlations between technical and biological covariates used in regression models to identify differentially expressed genes.** A) Comparison between AAA and control samples and B) Comparison between aneurysms of different diameter. Abbreviations: FC: flow cell type; Lane: flow cell lane; Library: date of creation of the library; percentage of RNA fragments (DV200); GC_mean: GC mean content; Date: date of creation of the library; Batch: batch number.

<

B

A
